# Incidence and Predictors of Mortality among Neonates Admitted for Congenital Heart Disease in Public Comprehensives Specialized Hospitals, Amhara Region, Ethiopia 2023

**DOI:** 10.1101/2023.09.24.23296057

**Authors:** Abatwoy Ayfokru, Sisay Shewasinad, Fuad Ahmed, Mitku Tefera, Genet Nigusie, Emawaysh Getaneh

**Affiliations:** Department of Nursing, school of nursing and Midwifery Asrat woldeyes health science campus, Debre Berhan University, Ethiopia; Department of Midwifery Debre-Birehan health Science College, Amhara region, Ethiopia; Department of Nursing Mida-woremo primary Hospital, Amhara region, Ethiopia; Department of Midwifery, school of nursing and Midwifery Asrat woldeyes health science campus, Debre Berhan University, Ethiopia

**Keywords:** Congenital Heart disease, Neonate, mortality. Incidence, predicators. Ethiopia

## Abstract

**Background:** Congenital heart disease is typically defined as a structural abnormality of the heart and great vessels present at birth. The incidence and mortality of congenital heart disease in neonates vary worldwide and it is the leading cause of birth defects, morbidity and mortality.

**Objective:** to assess incidence and predictors of mortality among neonates admitted for congenital heart disease in public comprehensives specialized hospitals Amhara region, Ethiopia a Retrospective follow-up study 2023

**Method:** Institution-based retrospective follow-up study was conducted among 583 neonates with congenital heart disease admitted to Amhara region public comprehensive specialized hospitals from January 1^st^ 2018 to December 30^th^, 2022. The study subjects was selected using simple random sampling techniques. Data were entered into EPI data version 4.6, then exported to STATA version 16. The log-rank test and Kaplan Meier failure curve was used to estimate and compare the probability of death. First bivariable then multivariable regression was done. After computing a hazard ratio with a 95% confidence interval, variables with a p-value of 0.05 or lower was considered to be significant predictors of mortality

**Results:** Out of 583 congenital heart disease neonate 9.9% were developed death. The incidence rate of mortality among congenital heart disease neonates was 11.9% per 1000 with 4844 person-days of observation (95% CI;-0.009-0.015). In this study were found to be Sepsis(AHR:2.24(95%CI:-1.184.23),Cyanoticcongenital heart disease(AHR:3.49(95%CI:1.93-6.28), Home delivery(AHR:1.9(95%,CI:1.06-3.6), had maternal history of Gestational diabetes multiuse (AHR: 1.94 (95% CI 1.04-3.61). Neonates having another congenital malformations. (AHR 2.49(1.33-4.67). neonates have been Airway resuscitation needed during delivery (AHR: 0.43(95%CI: 0.18-0.98). were found to be a significant predictors of Congenital heart disease neonatal mortality.

**Conclusion and recommendation:** The incidence rate of neonatal mortality among congenital heart disease neonate was high as compared to findings of other studies. sepsis, cyanotic congenital heart disease, home delivery, maternal history of gestational diabetes malts, others congenital malformation and Airway resuscitations needed during delivery were significant predictors of mortality for neonates with congenital heart disease. Hence, giving special attention to patients with identified predictors should be better

## Introduction

Congenital heart diseases (CHDs) are defined as a gross structural abnormality of heart or intrathoracic great vessels due to congenital anomalies. This can affect blood flows through the heart and the rest of the body.[1, 2] It is one of the most common congenital defects and accounts for nearly one-third of all major congenital anomalies[3].In 2016 The World Health Organization informed that 9,274,915 children died due to heart anomalies. Congenital heart disease can have genetic causes. But can also result after exposure to several risk factors (i.e., environmental factors, maternal health and age)

There are several common forms of CHDs. including Ventricular Septal Defect (VSD), Atrial Septal Defect (ASD), Patent Ductus Arteriosus (PDA), Coarctation of Aorta (CoA), Tetralogy of Fallot (TOF), Transposition of Great Arteries (TGA), Pulmonary Stenosis (PS), and Aortic Stenosis (AS) are common types of CHDs,[4, 5]

Atrial and ventricular septal defects accounted for 51% of cases, with a rise in incidence over time[5]. About 30–40% of CHD patients show symptoms in the first year of life, although 60% of them can be diagnosed by the first month of life.[6]

Congenital heart disease is related to increased neonatal death[7, 8].Approximately one-third of the congenital heart disease occurred during the first week of gestational age[9]. The mortality rate from congenital heart disease is higher in preterm newborns with low birth weight and comorbidity, the main cause of death was cardiogenic shock[10]. The neonatal mortality was still high during the first 28 day of life with congenital heart disease which indicates that continuous monitoring and early intervention may be beneficial[11]. Prevention and screening are key measures for lowering mortality rate among neonates with congenital heart disease. However, this is a difficult task. because public health workforce, epidemiologists, and pediatric heart surgeons are under staffed, poor health care infrastructure, lack of awareness to congenital heart disease[12, 13].

Globally over the last 15 years around 1.35 million live births are diagnosed with CHD every year [5].Worldwide 90% of newborns with CHD received inadequate medical care [14]. In 2017 the incidence rate of CHD was 17.9/1000 worldwide with 19.1/1000 for males and 16.6/1000 for females[15, 16]. Thirty percent of neonates die from CHD in the first month after birth[17]. According to the 2019 WHO report 217,000 neonates died due to CHD [18].Nine-tenths of the world’s babies born with CHD live in locations with little to no care where mortality remains high[3]. Every year an estimated 500,000 neonates in Africa born with CHD and need special treatment from a cardiology specialist. Congenital heart disease is a severe non-communicable disease that has a significant impact in low-and middle-income countries, the burden of congenital heart disease is vast, this major non communicable disease is largely hidden[19]

Asia, a population of 16 million people is served by one cardiac center[20]. There is one cardiac center for every 33 million individuals in Africa[21]. In Ethiopia one cardiac center served for 56 million people. Many of them died before receiving a diagnosis and the affected children’s families must travel significant distances for surgery and follow-up. CHD consequently has a negative impact on the daily activities and financial situation of family members that leading to decline in quality of life.in addition neonates with heart disease frequently have poor nutrition, infections, and pulmonary hypertension in low-income countries like Ethiopia which makes treatment more difficult. Overall providing care of these neonates is challenging since cardiac care requires a lot of resources, such as infrastructure, skilled personnel, and equipment.[22–25]. Congenital heart disease in neonates the second leading cause of death in the first year of life after infectious diseases[26].

Mortality in the first year of life was 12 times more likely in newborns born with severe forms of congenital heart disease[27],thus hundreds of neonates die each year from CHD, while millions more remain in desperate need of treatment in the developing countries[28] [29]. Neonatal mortality due to CHD is significantly underreported in sub-Saharan Africa[21, 23]. The targets for the Sustainable Development Goals and the Millennium Development Goals of reducing infant mortality are significantly affected by CHD [18].

Heart diseases require advanced diagnostic skills and delicate surgical procedures, which require a well-organized health system. Prior research shows that the Ethiopian health care system does not have enough medical personnel to diagnose CHD[30]. In addition to an inefficient health care system, the cost of preventing and treating heart diseases in Ethiopia poses a great burden on households[31]. Ethiopia About 66.7% of children with structural heart disease develop heart failure. It’s known that heart failure is a risk for neonatal mortality and the case fatality rate of HF was 13.9 %[32].

Estimating the scope of public health impact of congenital heart disease is significantly under estimated due to inadequate vital registration systems and inaccurate records of cause of death in developing countries. the recommendation of the International Society of Ultrasound and Gynecology to include the four-chamber view and visualization of the outflow tract views in the routine prenatal ultrasound assessment of the fetal heart is supported by evidence and has improved the rate of prenatal CHD detection and decreased mortality rates[33]. Preconception care decrease burden of occurrence of congenital malformation and malformation related mortality[34].

Similarly as far as the investigator searches there is limited evidence on the incidence and predicators of mortality among neonates with congenital heart disease in Ethiopia. Thus, this study investigated the incidence and predicator of mortality among neonates with congenital heart disease admitted to public comprehensives specialized Hospitals. Amhara region Ethiopia 2023. Therefore, the objective of this study was to assess incidence and predictors of mortality among neonates with congenital heart disease in public comprehensives specialized hospitals, amhara region, in Ethiopia (show fig 1).

**Figure 1.**
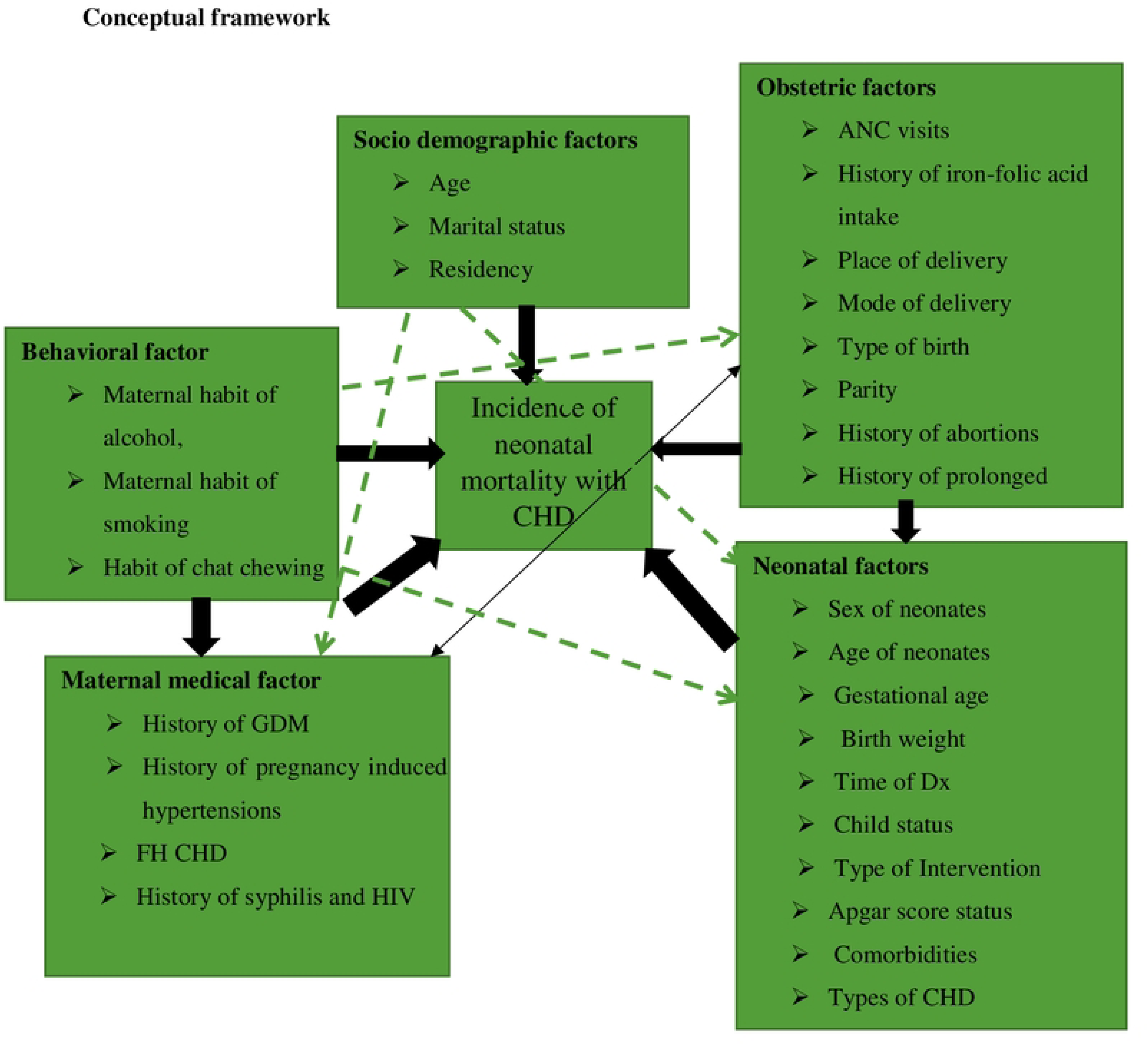
Schematic presentation of conceptual framework of incidence and predictors of mortality among neonates admitted for congenital heart disease in public comprehensives specialized hospitals, amhara region in Ethiopia 2023.

## Method

### Study area and period

The study was conducted from April 01 to 15/08 /2015, at selected governmental Comprehensive Specialized Hospitals in the Amhara regional state in Ethiopia. The capital city of Amhara region is Bahir Dar which is the seat of the Regional Health bureau (ARHB) of Amhara,

According to 2020 the Health and Health Related Indicators published by MoH. Amhara has 82 Hospitals, 861 Health Centers and 3,565 Health Posts. Among those 82 hospitals in the region, the University of Gondar, Dessie, Felege-Hiwot, Tibebe-Ghion, Debre Markos, Woldia, Debre Tabor, and Debre Berhan are Comprehensive Specialized Hospitals.

### Study design

Institutional based retrospective follow-up study was conducted

### Source population

All neonate with congenital heart disease and admitted to the NICU wards of Amhara regional state Comprehensive Specialized Hospitals, From January 1^st^ 2018 to December 30^th^ 2022, were the source population.

### Study population

Selected neonates with congenital heart disease were admitted to the NICU wards of Amhara regional state’s Selected Comprehensive Specialized Hospitals, from January 1^st^ 2018 to December 30^th^ 2022.

### Eligibility criteria

#### Inclusion criteria

All newly diagnosed and admitted neonates with congenital heart disease from January 1^st^ 2018 to December 30^th^ 2022.

### Exclusion criteria

Congenital heart disease neonates with incomplete records (not registered date of admission, date discharge, neonatal status, and neonatal age), and unknown outcome status were excluded the study.

### Sample size determination

For the first specific objectives sample size was determined by using single population proportion formula by considering the following assumptions. Prevalence (p) of 35.8% taken from a study in Addis Ababa referral public hospitals[35] 95% confidence level, 5% margin of error and 10% of incomplete records.

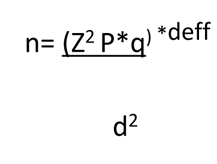

Where; p was the proportion of prevalence and associated factor of congenital heart disease among congenital anomalies (0.358%) d=5% or 0.05 because the confidence interval is 95%

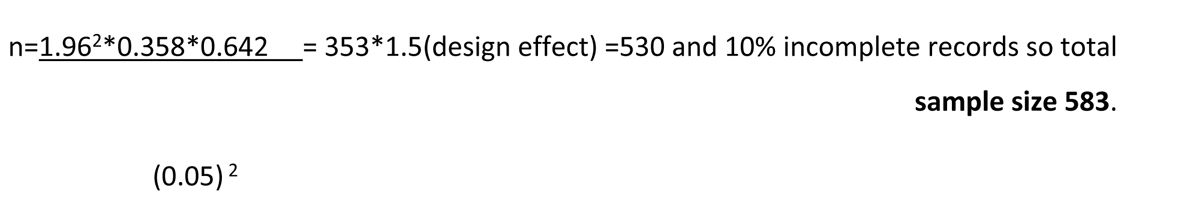

### Sampling technique and Sampling procedure

For comprehensive specialized hospitals were selected randomly by lottery method. Number of participants to each Hospital was allocated by proportion to population size. The sampling interval of each hospital were calculated by dividing the number of eligible study participants for allocated sample size of that Hospital, finally the first participants were selected by using systematic random sampling technique every K^th^ (7176/583=12) intervals. The first participants were selected by lottery method from 1-12 and the number 6 was selected. Then, the next selection was continued every 12^th^ intervals until obtained the required sample size was achieved (Show fig 2).

**Figure 2.**
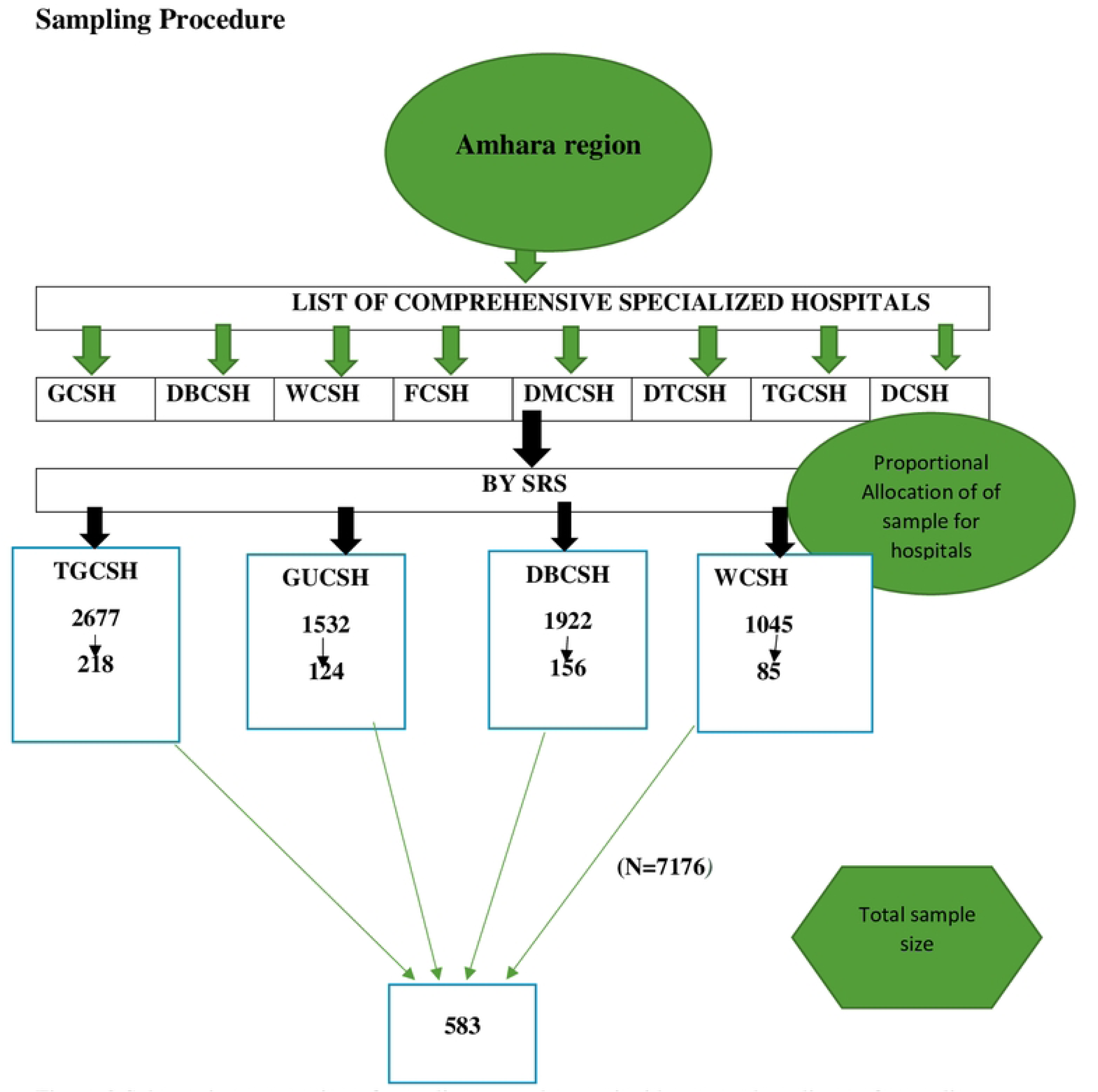
Schematic presentation of sampling procedure on incidences and predictor of mortality among neonates with congenital heart disease at Amhara region public Comprehensive Specialized Hospitals, Ethiopia, 2023.

### Study variables Dependent Variables

- Incidence of neonatal mortality with CHD

#### Independent variables

**Socio-demographic factors:** Maternal age, marital status, Residency

**Maternal behavioral factors**: Maternal habit of alcohol intake, smoking, chat chewing,

**Maternal medical factors:** History of maternal DM. pregnancy induced hypertension, HIV, cardiac problems, TB, anemia, Syphilis, Family history of CHD

**Maternal obstetrical factors:** ANC visits. History of folic acid intake, Place of delivery, Mode of delivery, history of Abortion, Type of birth, gravidity, parity. Preterm delivery, RH factors, prolonged delivery

**Neonatal factors:** Age of neonates, sex of neonates, Birth weight. Apgar score status, Gestational age, date of admission and date of discharge, types of intervention,, comorbidities, length of hospital stay, types of CHD, child status and other congenital anomalies

### Data collection tool and procedure

A pretested pre-structured checklist were collect data from the NICU registration book. The checklist for data extraction tool consists of socio demographic related variables, maternal behavioral and medical related variables, obstetric and neonatal related variables, Time from the admission of CHD was the starting point for retrospective follow-up and the endpoint was the date of discharge. The recordings of all study participants were selected according to the eligibility criteria. The questionnaire was adapted and modified from reviewing different literature and guidelines[10, 29, 35–41], The questionnaire was prepared as a checklist in the English language, The reliability of the tool were assessed by conducting a pretest study and face and content validity was ensured, to ensure its eligible and to avoid any confusion regarding the understanding of the questionnaire and the questionnaire was(occupational status, educational status, History of taking of medication during pregnancy, History of mal-presentation, heart auscultation status, types of CHD, time of diagnosis to CHD) modified based on feedback from the study, Health professionals who have been working in the treatment center were collected the data. Prior to data collection period, one day of training was given for the data collectors and supervisors on the objectives of the study, data collection method and tools and how to maintain the confidentiality of the data. During data collection time, Supervisors and data collectors was daily discussed, on faced problems during data collection time until the data collection was accomplished.

### Operational definitions

**Neonate**: The first 28 days of life[42]

**Censored;** refers to neonates with congenital heart disease were improved, discharged, or against medical treatment, refers to others center and outcome known after 28 days

**Event;** - The event was death of neonates with congenital heart disease while in the hospital and whose death report was recorded with in 28days.

**Time to event; -** A time from the admission date by CHD to the discharge date while the neonate is event. It will measured by subtracting the date of admission from the date of discharge

**Co-morbidities;** Newborns with CHD and have other illnesses such as sepsis and under nutrition [43]

**Incomplete charts**: when one of the independent variables was not registered (date of admission and discharge, discharge status, and neonatal age)

**Medical disorders in the mother**: - Any registered medical diagnosis history of the mother on the maternal and neonate’s medical record[44]

**Antenatal care visit**: - Any history of follow up care provided by skilled health-care professionals for both mother and baby during pregnancy designated or recorded on chart[44]

**Low APGAR SCORE: -** A neonate who have an Apgar score of <7[45]

**Preterm (premature) birth:** - Neonate born before 37 weeks of gestational age[45]

**Low birth weight: -** neonates who have <2500 gram of birth weight[46]

**Congenital heart defect; -**major or minor congenital anomalies defined as anatomical structural and functional defect present at birth which was confirmed by pediatricians or echocardiography [47]

### Data quality control assurance plan

The checklist was pre-tested among 5% of the study participants in other than the study area (Dessie Referral Hospital) to assure the data quality. The necessary modifications and corrections were made to standardize and ensure its validity. Experts in the clinical and academic area were involved to evaluate the content of the questionnaire. The tool utilized in the current study was validated and found to be reliable. The data collectors and supervisor were senior experienced BSc nurses who are working in NICU (one supervisor and two data collectors were select each hospital). One day of training was given on the purpose of the study, the data collection tool (checklist), data collection methods, and ethical concerns during data collection. The supervisor was monitored the data collection process. The principal investigator and supervisor have done spot-checking and reviewing the complete checklist by the data collectors to ensure the completeness and consistency of the collected information (data). Consistency was examined through a random selection of cards by the principal investigator, during the data collection time, close supervision and monitoring were carried out by the supervisors and data entry were also done using Epi Data software.

### Data Processing and Analysis

After checking data completeness and consistency, the collected data was coded and entered into Epi- data version 4.6 then exported into STATA version 16 for cleaning and analysis. Data were summarized with text, table, figure, mean, and SD based on the data nature. The frequency distribution was used for categorical data and results were presented as graphs & tables. During the analysis, the outcome was used as a dependent variable indicating death as an event of interest. The outcome of each participant was dichotomized into death (event of interest), coded as “1”, and censored coded as “0”. A variance inflation factor (VIF 1.06) to assess multi-collinearity between independent variables). Was done. “Kaplan-Meier survival estimates” was used to estimate the mean survival time, cumulative probability of survival and survival differences between covariates were done. A log-rank test was used to assess statistically differences across groups of independent variables. The proportionality assumptions were tested using a Schoenfeld residual global test (p value 0.6337).a Cox-Snell residual test was used to evaluate the model’s goodness of fit. in order to identify predictors of mortalities, A bivariable and multivariable Weibull regression models was used. Variables with a P-value of less than 0.25 in the bivariate analysis was entered into the multivariate analysis. In multivariable analysis, a variable with a p-value less than 0.05 at a 95% confidence level was considered as an independent predictor of mortality in CHD neonates.

### Ethical consideration

The IRB board of Debre Berhan University’s Asrat Woldeys Health Science Campus granted ethical clearance (protocol **number= IRB 123**). For data collection, the University submitted a formal letter of cooperation to the selected public hospital administrative body. Permission was obtained from all relevant bodies at the chosen hospital. The data collectors were also in charge of searching for and obtaining the medical records of the chosen samples. The confidentiality of patients’ records was maintained by omitting their names and personal identification from the data collection format.

## Results

### Socio-demographic characteristics of mother of neonate

A total of 583 neonates with CHD medical charts were reviewed, of which 566 were complete the remaining 11 were incomplete, and 6 weren’t available at the time of data collection. However, these charts were replaced with other charts by randomly selecting charts that hadn’t been chosen earlier. In this study from the total study participants 525 (90.5%) were censored and 58 (9.95%) were events (death), the mean age of participants was 32.80 (SD ± 0.30) years (95% CI 32.20--33.40) Over half of the mothers, 310 (53.17%) were older than 35 years. Nearly 506 neonatal mothers (86.79%) were married. About 69.3 % of the neonates had age greater than seven days. Three hundred ninety three participants (67.41%) were females. More than three fourth of participants 487 (83.53%) were rural residence (Table 1).

**Table1:** Socio demographic characteristics predictor’s of neonatal mortality with congenital heart disease admitted in NICU ward of public comprehensives specialized hospitals Amhara region, Ethiopia 2023 retrospective follow up study *from Jannuary1, 2018-December 30, 2022*.

| Covariates | Category | Frequency (%) | Status |  |
| --- | --- | --- | --- | --- |
|  |  |  | Censored (%) | Death (%) |
| Maternal age | <= 20 Year | 37(6.35%) | 36(6.86%) | 1(1.72 %) |
|  | 20=34 Year | 236(40.48%) | 215(40.95%) | 21(36.21%) |
|  | >=35 Year | 310(53.17%) | 274(52.19%) | 36(62.07) |
| Sex of neonates | Male | 190(32.59%) | 168(32%) | 22(37.93%) |
|  | Female | 393(67.41%) | 357(68%) | 36(62.07%) |
| Age of neonates | <=7 day | 179((30.70%) | 146(27.81%) | 33(56.90 %) |
|  | >=8 day | 404(69.30) | 379(72.19%) | 25(43.10 %) |
| Marital Status | Married | 506(86.79%) | 459(87.43%) | 47(81.03%) |
|  | Single | 13(2.23%) | 10(1.90%) | 3(5.17%) |
|  | Divorced | 41(7.03%) | 34(6.48%) | 7(12.07%) |
|  | Widowed | 23(3.95%) | 22(4.19%) | 1(1.72%) |
| Residences | Urban | 96(16.47%) | 77(14.67%) | 19(32.76%) |
|  | Rural | 487(83.53%) | 448(85.33%) | 39(67.24%) |

### Maternal behavioral factors

Among 583 neonatal mothers participated in this study, 106 (18.18%) of them drank alcohol, 135 (23.16%) smoked cigarettes, and 118 (20.24%) chewed khat during this pregnancy (Table 2).

**Table 2:** Maternal behavioral factors characteristics of predictor’s of neonatal mortality with congenital heart disease admitted in NICU ward of public comprehensives specialized hospitals Amhara region, Ethiopia 2023 retrospective follow up study from Jannuary1, 2018-December 30, 2022.

| Covariates | Category | Frequency (%) | Status |  |
| --- | --- | --- | --- | --- |
|  |  |  | Censored (%) | Death (%) |
| History of Alcohol intake during Pregnancy | Yes | 106(18.18%) | 85(16.19%) | 21(36.21%) |
|  | No | 477(81.82%) | 440(83.81%) | 37(63.79%) |
| History of Smoking intake during this pregnancy | Yes | 135(23.16%) | 127(24.19%) | 8(13.79%) |
|  | No | 448(76.84%) | 398(75.81%) | 50(86.21%) |
| History of Chewing chat during this pregnancy | Yes | 118(20.24%) | 114(21.71%) | 4(6.90%) |
|  | No | 465(79.76%) | 411(78.29%) | 54(93.10%) |

### Maternal Medical Factors

Of the total neonatal mothers included in this study, 72 (13.35%) had gestational diabetes mellitus, 40 (6.66%) had cardiac problems, 57 (9.78%) had HIV/AIDS, 37 (6.35%) had syphilis, and 129 (22.13%) had a history of congenital heart disease (CHD), 22 (3.77%) of mothers had a history of TB, and 17 (2.92%) of mothers had a history of hypertension (Table 3).

**Table 3:** Maternal medical characteristics of predictor’s of neonatal mortality with congenital heart disease admitted in NICU ward of public comprehensives specialized hospitals Amhara region, Ethiopia 2023 retrospective follow up study from Jannuary1, 2018-December 30, 2022.

| Covariates | Category | Frequency (%) | Status |  |
| --- | --- | --- | --- | --- |
|  |  |  | Censored (%) | Death (%) |
| GDM | Yes | 72(13.35%) | 52(9.90%) | 20(34.48%) |
|  | No | 511(87.65%) | 473(90.10%) | 38(65.52%) |
| Hypertensions | Yes | 17(2.92%) | 16(3.05%) | 1(1.72%) |
|  | No | 566(97.08%) | 509(96.95%) | 57(98.28%) |
| Cardiac problems | Yes | 40(6.86%) | 34(6.48%) | 6(10.34%) |
|  | No | 543(93.14%) | 491(93.52%) | 52(89.66%) |
| Syphilis | Yes | 37(6.35%) | 30(5.71%) | 7(12.07%) |
|  | No | 546(93.65%) | 495(94.29%) | 51(87.93%) |
| HIV/AIDS | Yes | 57(9.78%) | 37(7.05%) | 20(34.48%) |
|  | No | 526(90.22%) | 488(92.95%) | 38(65.52%) |
| TB | Yes | 22(3.77%) | 17(3.24%) | 5(8.62%) |
|  | No | 561(96.23%) | 508(96.76%) | 53(91.38%) |
| Anemia | Yes | 38(6.52%) | 33(6.29%) | 5(8.62%) |
|  | No | 545(93.48%) | 492(93.71%) | 53(91.38%) |
| History of CHD | Yes | 129(22.13%) | 110(20.95%) | 19(32.76%) |
|  | No | 454(77.87%) | 415(79.05%) | 39(67.24%) |

### Maternal obstetrical factors

The majority of neonatal mothers, 506 (86.79%) had two or more pregnancies, and 452 (77.53%) had spontaneous vaginal births and 301 (51.63%) had ANC follow-up (Table 4)

**Table 4.** Maternal obstetrical factors characteristics of predictor’s of neonatal mortality with congenital heart disease admitted in NICU ward of public comprehensives specialized hospitals Amhara region, Ethiopia 2023 retrospective follow up study from Jannuary1, 2018-December 30, 2022.

| Covariates | Category | Frequency (%) | Status |  |
| --- | --- | --- | --- | --- |
|  |  |  | Censored (%) | Death (%) |
| ANC follow Up visits | Yes | 301(51.63%) | 266(50.67%) | 35(60.34%) |
|  | No | 282(48.34%) | 259(49.33%) | 23(39.66%) |
| Ferrous sulphate with folic acid provided | Yes | 305(52.35%) | 283(53.90%) | 22(37.93%) |
|  | No | 278(47.68%) | 242(46.10%) | 36(62.07%) |
| Obstetric U/S performed within 24 WK of GA | Yes | 129(22.13%) | 116(22.10%) | 13(22.40%) |
|  | No | 454(77.87%) | 409(77.90%) | 45(77.59%) |
| Gravidity | Premi gravida | 77(13.21%) | 70(13.33%) | 7(12.07%) |
|  | Multi gravida | 506(86.79%) | 455(86.67%) | 51(87.93%) |
| History of Abortion | Yes | 106(18.18%) | 89(16.95%) | 17(29.31%) |
|  | No | 477(81.82%) | 436(83.05%) | 41(70.69%) |
| History of neonatal death | Yes | 108(18.52%) | 90(17.14%) | 18(31.03%) |
|  | No | 475(81.48%) | 435(82.86%) | 40(68.97%) |
| History of APH | Yes | 43(7.38%) | 36(6.86%) | 7(12.07%) |
|  | No | 540(92.62%) | 489(93.14%) | 51(87.93%) |
| History of prolonged labour | Yes | 44(7.55%) | 38(7.24%) | 6(10.34%) |
|  | No | 539(92.45%) | 487(92.76%) | 52(89.66%) |
| History of Premature Rapture of membranes | Yes | 44(7.55%) | 32(6.10%) | 12(20.69%) |
|  | No | 539(92.45%) | 493(93.90%) | 46(79.31%) |
| History of RH Factors | Yes | 55(9.43%) | 49(9.33%) | 6(10.34%) |
|  | No | 528(90.57%) | 476(90.67%) | 52(89.66%) |
| Types of birth | Single | 270(46.31%) | 252(48%) | 18(31.03%) |
|  | multiple | 313(53.69%) | 273(52%) | 40(68.97%) |
| Mode of delivery | SVD | 452(77.53%) | 409(77.90%) | 43(74.14%) |
|  | C/S | 131(22.47%) | 116(22.10%) | 15(25.86%) |
| Place of delivery | Home | 165(28.30%) | 133(25.33%) | 32(55.17%) |
|  | Health center | 155(26.59%) | 148(28.19%) | 7(12.07%) |
|  | Hospital | 263(45.11%) | 244(46.48%) | 19(32.76%) |

### Neonatal related characteristics

Among newborns with CHD who were admitted to the NICU, 389 (66.72%) had low birth weight, 260 (44.60%) had neonatal sepsis, 172 (29.50%) had PNA, 320 (54.89%) had preterm delivery, and 49 (8.10%) had an additional cardiac abnormality. At admission, the mean neonatal birth weight was 2253 *grams (Table 5)*.

**Table 5:** Neonatal medical & other characteristics of CHD neonates admitted to NICU at Amhara region public comprehensives specialized hospitals from Jannuary1, 2018-December 30, 2022.

| Covariates | Category | Frequency (%) | Status |  |
| --- | --- | --- | --- | --- |
|  |  |  | Censored (%) | Death (%) |
| Gestational age | <=37 WK | 320(54.89%) | 285(54.29%) | 35(60.34%) |
|  | >=37WK | 263(45.11%) | 240(45.71%) | 23(39.66%) |
| 5<sup>th</sup> minutes Apgar score Status | Normal | 40(6.86%) | 39(7.43%) | 1(1.72%) |
|  | Moderate | 313(53.69%) | 296(56.38%) | 17(29.31%) |
|  | Severs | 230(39.45%) | 190(36.19%) | 40(68.97%) |
| Birth weight | <=2500 | 389(66.72%) | 351(66.86%) | 38(65.52%) |
|  | >=2500 | 194(33.28%) | 174(33.14%) | 20(34.48%) |
| Time of Diagnosis to CHD | Early diagnosis | 217(37.22%) | 192(36.57%) | 25(43.10%) |
|  | Late diagnosis | 366(62.78%) | 333(63.43%) | 33(56.90) |
| Types of CHD | Cyanotic CHD | 117(20.07%) | 84(16.00%) | 33(56.90%) |
|  | Asyanotic CHD | 466(79.93%) | 441(84.00%) | 25(43.10%) |
| Others congenital malformation | Yes | 49(8.40%) | 35(6.67%) | 14(24.14%) |
|  | No | 534(91.60%) | 490(93.33%) | 44(75.86%) |
| Comorbidity | Yes | 291(49.91%) | 250(47.62%) | 41(70.69%) |
|  | No | 292(50.09%) | 275(52.38%) | 17(29.31%) |
| Types of intervention to CHD | Medical | 514(88.16%) | 459(87.43%) | 55(94.83%) |
|  | Surgical | 69(11.84%) | 66(12.57%) | 3(5.17%) |
| Air way Resuscitation needed during delivery | Yes | 374(64.15%) | 323(61.52%) | 51(87.93%) |
|  | No | 209(35.85%) | 202(38.48%) | 7(12.07%) |
| PNA | Yes | 172(29.50%) | 165(31.43%) | 7(12.07%) |
|  | No | 411(70.50%) | 360(68.57%) | 51(87.93%) |
| Sepsis | Yes | 260(44.60%) | 217(41.33%) | 43(74.14%) |
|  | No | 323(55.40%) | 308(58.67%) | 15(25.86) |
| SAM | Yes | 133(22.81%) | 108(20.57%) | 25(43.10%) |
|  | No | 450(77.19%) | 417(79.43%) | 33(56.90%) |

### The incidence rate of mortality among congenital heart disease neonates

A total of 583 CHD neonates that were admitted to NICU had been followed from 1 to 28 days. The Overall mean hospital stay was 8.64 (95%CI: 8.17-9.12) with a minimum and maximum follow-up time of 1 and 28 days and the total person-time at risk was 4844 person day observations, In this finding, 58(9.79%) of the study participant were events (death) from 583 CHD neonates, 525 (90.05%) were censored (recovered, left NICU against medical advice, outcome after 28 days and referred to other center) at the end of the follow-up (Figure 3).

**Figure 3.**
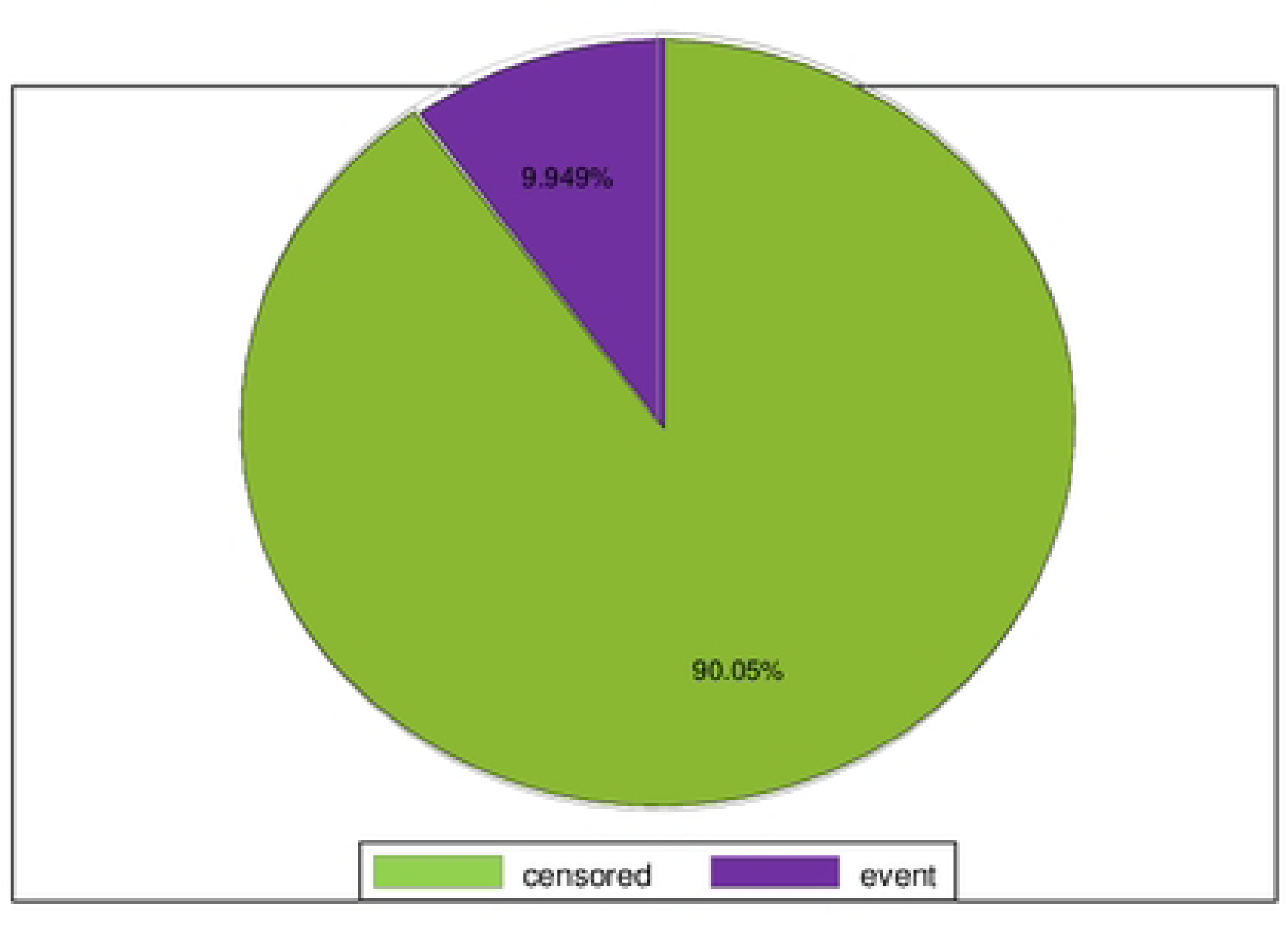
Outcome of neonates admitted to NICU of Amhara region comprehensives specialized hospitals in Ethiopia, January 1st, 2018 – December 31th, 2022.

The overall incidence rate of mortality among congenital heart disease neonates was 11.9 (95% CI;-0.009-0.015) per 1000 person days observation. In this study, the mortality rate among male and female congenital heart disease neonates was 13.1 and 11.3 per 1000 person day observations respectively. The rates of mortality among urban and rural residents were 21.3 and 9.8 per 1000 person day observations respectively

Early neonates (<7 days) had a higher rate of mortality 22.0 (95%% CI-0.015-0.031) as compared Late neonates ones (>=8 days) 7.4(95%;-CI.005-0.011) per 1000 person day observations. The mortality rate of congenital heart disease neonates varied across different categories of clinical characteristics of neonates. The incidence of mortality among patients presented with and without comorbidity was 16.5 and 7.2 per 1000 person day observations respectively. In addition, patients who had neonatal sepsis at admission revealed a higher incidence of mortality rate (21.0) than their counterparts (5.3) and sever acute malnutrition (23.0) without sever acute malnutrition (8.7) respectively.

The incidence rate of mortality of cyanotic CHD neonates (37.3) and Asyanotic CHD (6.3) and history of HIV/AIDS (41.4) without HIV/AIDS (8.7) per 1000 person day observations respectively.

This shows that neonates with maternal history of HIV/AIDS, cyanotic CHD, and sever acute malnutrition having the highest incidence of neonatal mortality with congenital heart disease than the above predictor variable

### Overall survival of congenital heart disease neonates admitted to NICU

The overall Kaplan-Meier estimate showed that the probability of survival of CHD neonates was high on the 1st day of admission, which comparatively decreases as follow-up time increases (fig 4).

**Figure 4.**
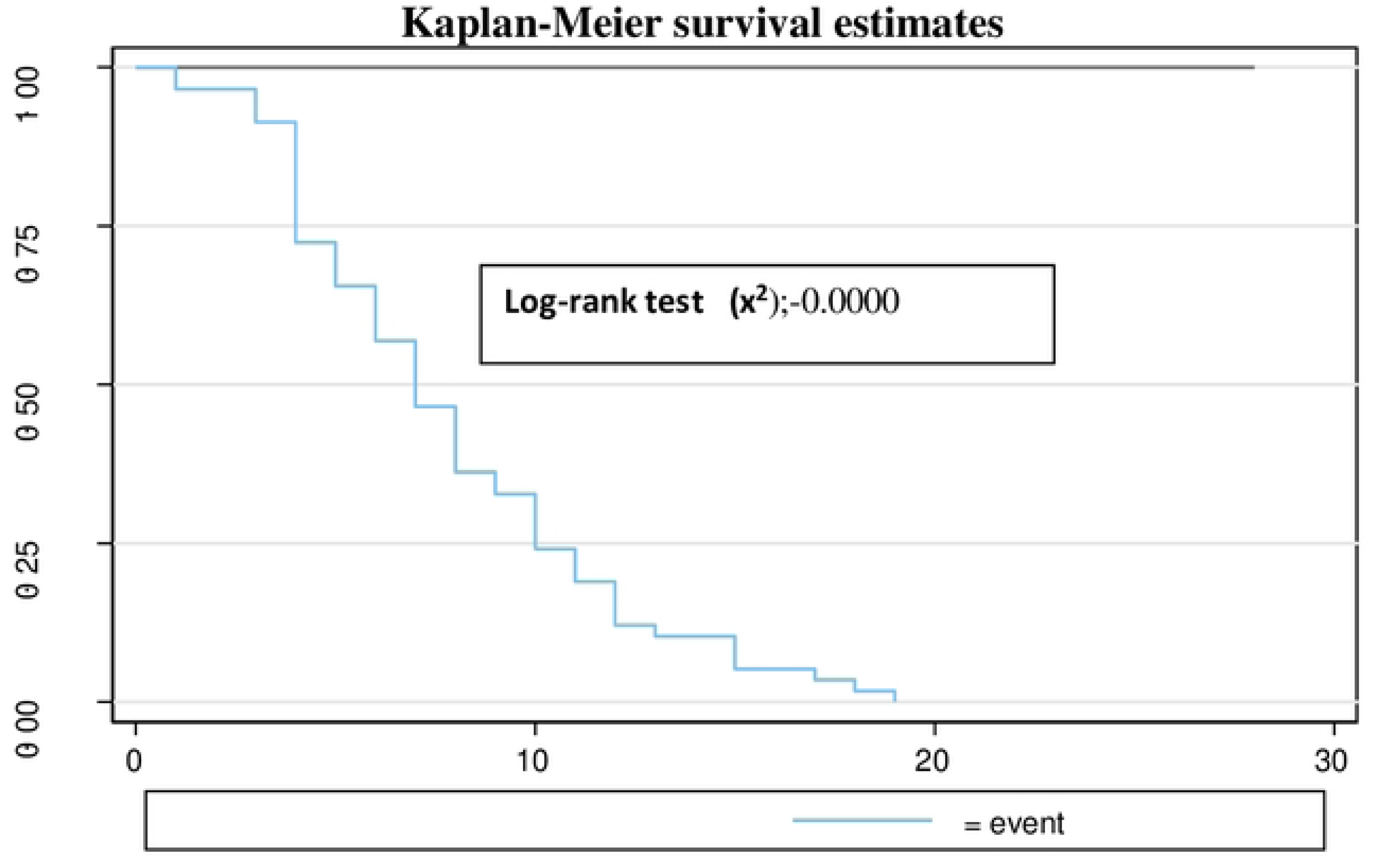
Overall Kaplan-Meir survival estimate of CHD neonates admitted to NICU at Amhara region comprehensives specialized public Hospitals in Ethiopia Jannuary1, 2018-December 30, 2022.

During the 1st day of hospital stay, a maximum 99.6% (95%CI: 98, 99.9) probability of survival was observed with a SD± of 0.0025. The overall mean survival time of neonates admitted to NICU in the study was 8 days with 90.3% (95% CI 0.86-0.93) a SD± 0.015. This study also showed that the probability of neonatal survival at the 7th and 14th day of hospital stay was 92.5% (95% CI 89 94) SD± = 0.013) and 82.1 % (95% CI: 76--86) SD± = 0.025), respectively. At the 20 days of hospital stay, the overall survival probability of neonates was 74.1% (95% CI: 65-80) with a standard error of 0.039 (Table 6).

**Table 6.** A life table showing survival probability of CHD Neonates was admitted to NICU selected public hospitals Amhara Region, Ethiopia, January 1st, 2018-December 30th, 2022.

| Interval |  | Beg. Total | Deaths | censored | Survival | Std. Error | [95% Conf. Int.] |
| --- | --- | --- | --- | --- | --- | --- | --- |
| 1 | 2 | 583 | 2 | 19 | 0.9965 | 0.0025 | 0.9861- 0.9991 |
| 2 | 3 | 562 | 0 | 33 | 0.9965 | 0.0025 | 0.9861 -0.9991 |
| 3 | 4 | 529 | 3 | 31 | 0.9907 | 0.0041 | 0.9778 -0.9961 |
| 4 | 5 | 495 | 11 | 51 | 0.9675 | 0.0080 | 0.9474 -0.9800 |
| 5 | 6 | 433 | 4 | 43 | 0.9581 | 0.0092 | 0.9357 -0.9728 |
| 6 | 7 | 386 | 5 | 60 | 0.9446 | 0.0109 | 0.9188- 0.9624 |
| 7 | 8 | 321 | 6 | 42 | 0.9257 | 0.0131 | 0.8953- 0.9476 |
| 8 | 9 | 273 | 6 | 36 | 0.9039 | 0.0155 | 0.8685- 0.9302 |
| 9 | 10 | 231 | 2 | 28 | 0.8956 | 0.0165 | 0.8583- 0.9236 |
| 10 | 11 | 201 | 5 | 26 | 0.8718 | 0.0192 | 0.8288 -0.9046 |
| 11 | 12 | 170 | 3 | 29 | 0.8550 | 0.0211 | 0.8078- 0.8913 |
| 12 | 13 | 138 | 4 | 15 | 0.8288 | 0.0242 | 0.7752- 0.8707 |
| 13 | 14 | 119 | 1 | 20 | 0.8212 | 0.0251 | 0.7655- 0.8648 |
| 14 | 15 | 98 | 0 | 17 | 0.8212 | 0.0251 | 0.7655- 0.8648 |
| 15 | 16 | 81 | 3 | 8 | 0.7892 | 0.0302 | 0.7225- 0.8416 |
| 16 | 17 | 70 | 0 | 13 | 0.7892 | 0.0302 | 0.7225- 0.8416 |
| 17 | 18 | 57 | 1 | 5 | 0.7747 | 0.0329 | 0.7020 -0.8318 |
| 18 | 19 | 51 | 2 | 5 | 0.7587 | 0.0359 | 0.6795- 0.8209 |
| 19 | 20 | 45 | 0 | 3 | 0.7413 | 0.0391 | 0.6552 -0.8090 |
| 20 | 21 | 41 | 0 | 3 | 0.7413 | 0.0391 | 0.6552 -0.8090 |
| 21 | 22 | 38 | 0 | 4 | 0.7413 | 0.0391 | 0.6552- 0.8090 |
| 22 | 23 | 34 | 0 | 5 | 0.7413 | 0.0391 | 0.6552 -0.8090 |
| 23 | 24 | 29 | 0 | 8 | 0.7413 | 0.0391 | 0.6552- 0.8090 |
| 24 | 25 | 21 | 0 | 7 | 0.7413 | 0.0391 | 0.6552- 0.8090 |
| 25 | 26 | 14 | 0 | 4 | 0.7413 | 0.0391 | 0.6552- 0.8090 |
| 26 | 27 | 10 | 0 | 3 | 0.7413 | 0.0391 | 0.6552 -0.8090 |
| 27 | 28 | 7 | 0 | 3 | 0.7413 | 0.0391 | 0.6552- 0.8090 |
| 28 | 29 | 4 | 0 | 4 | 0.7413 | 0.0391 | 0.6552- 0.8090 |

### Survival function among predictor variables

In this study, CHD Neonates with comorbidity had a lower survival time of 15 days (95% CI 73-85 :) of mean survival time compared to those who had not been diagnosed with comorbidity. At the 28 days of hospital stay, the overall survival of neonates with comorbidity and without comorbidity was 80 % and 92 %, respectively (Figure 5). This survival time difference was statistically significant with a Log-rank test (x^2^) p-value = 0.0021.

**Figure 5.**
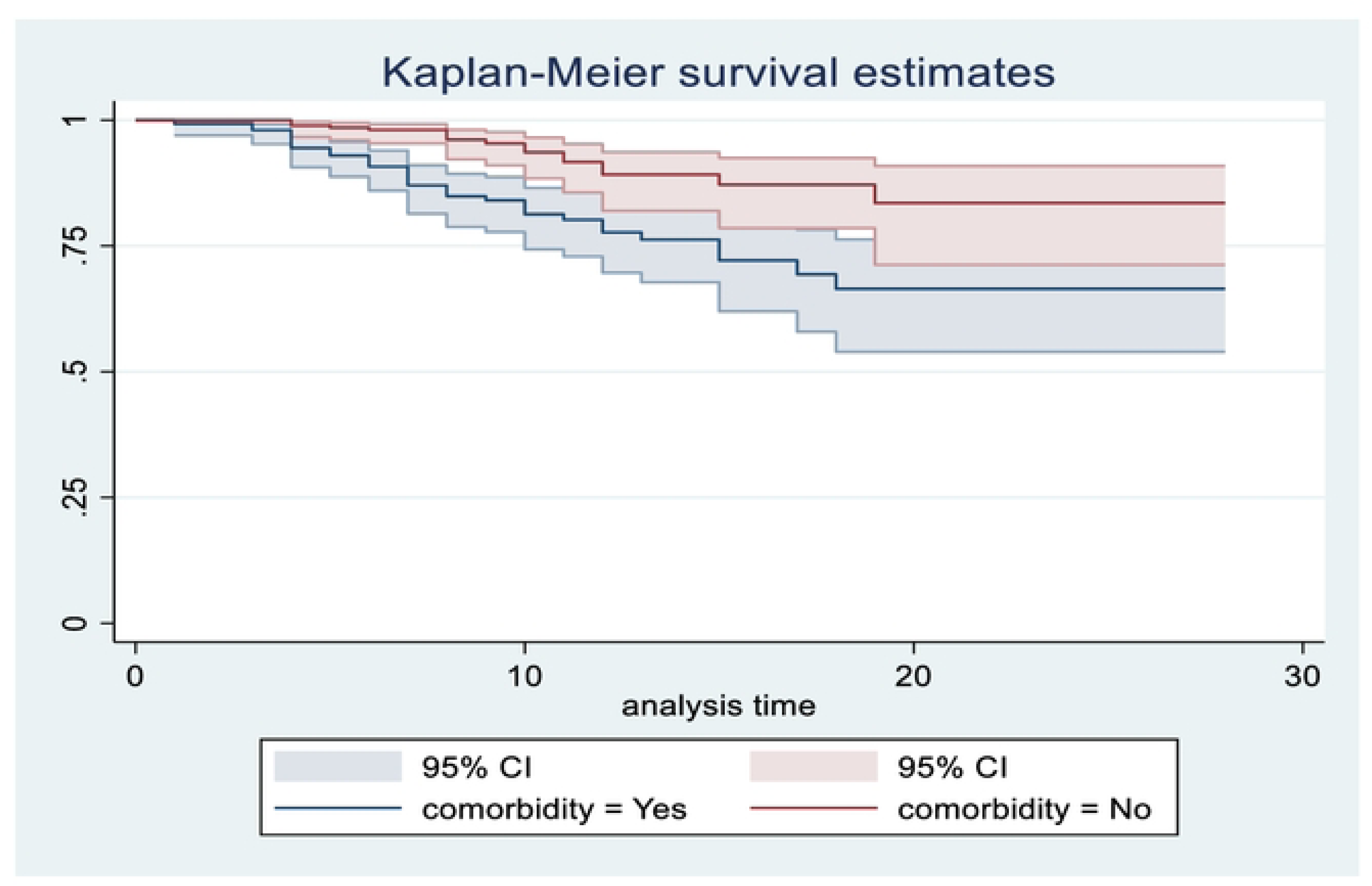
Kaplan-Meir survival estimate among CHD neonates admitted to NICU at public hospitals in Amhara Region with the category of comorbidity and without comorbidity from January 1, 2018-December 30^th^, 2022.

In this study, CHD Neonates with Cyanotic CHD had a lower survival time of 19 days (95% CI-67-81) of mean survival time compared to those who had been diagnosed with Asyanotic CHD. At the 28 days of hospital stay, the overall survival of neonates with Cyanotic CHD and Asyanotic CHD was 75.4% and 83%, respectively (Figure 6).

**Figure 6.**
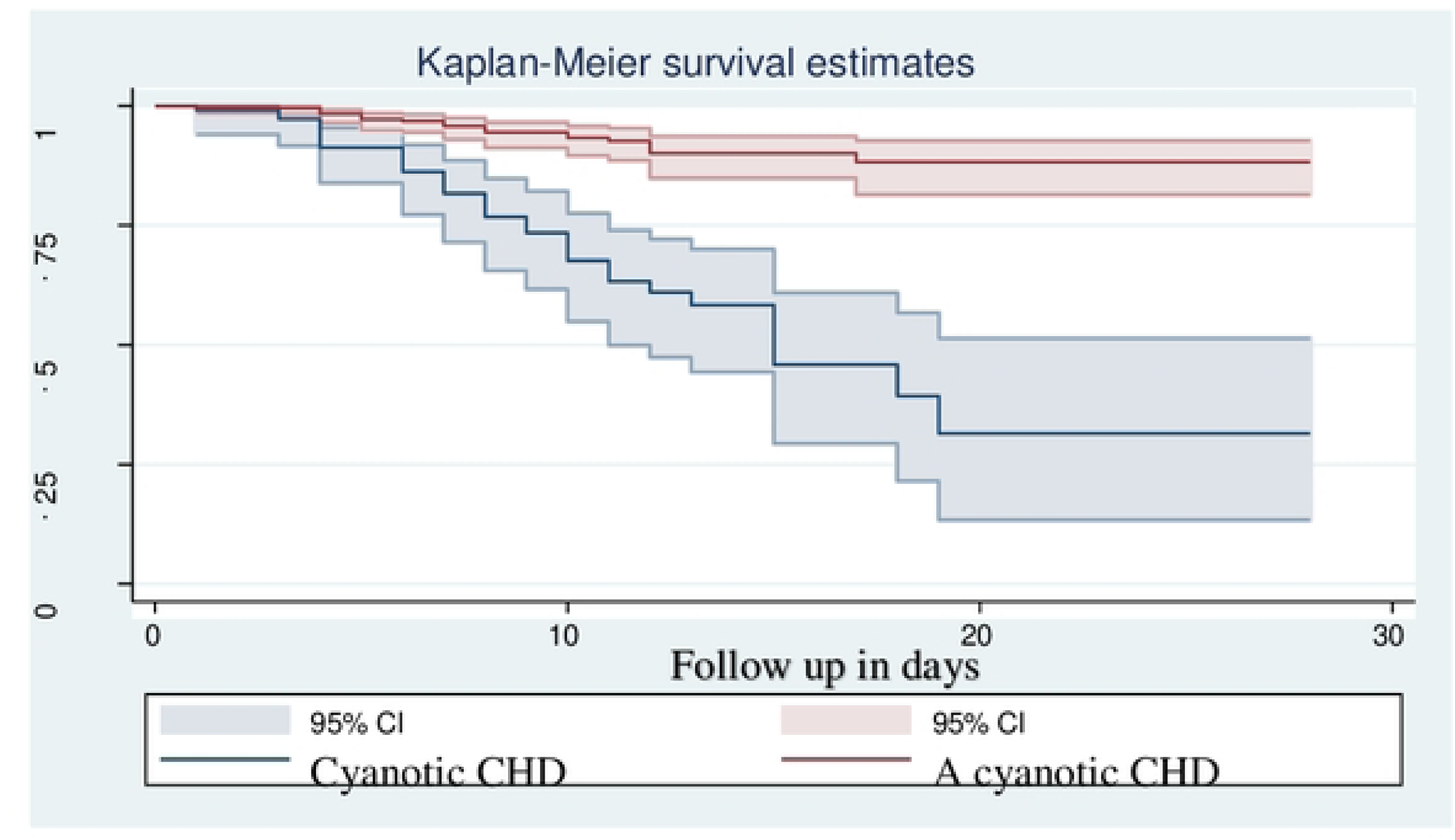
Kaplan-Meir survival estimate among CHD neonates admitted to NICU at public hospitals in Amhara Region with the category of Cyanotic and Asyanotic congenital heart disease from January 1, 20118-December 30, 2022.

As shown from the figure plotted below, the Kaplan Maier survival function for CHD neonates from mothers who have HIV positive was, a lower survival time (median survival time of 18 days, 95% than those Who have no HIV (no median survival time with overall survival probability of 77% and 83% % at the end of the study period respectively. The difference was statistically significant at a p-value = 0.000 (Show figure 7).

**Figure 7.**
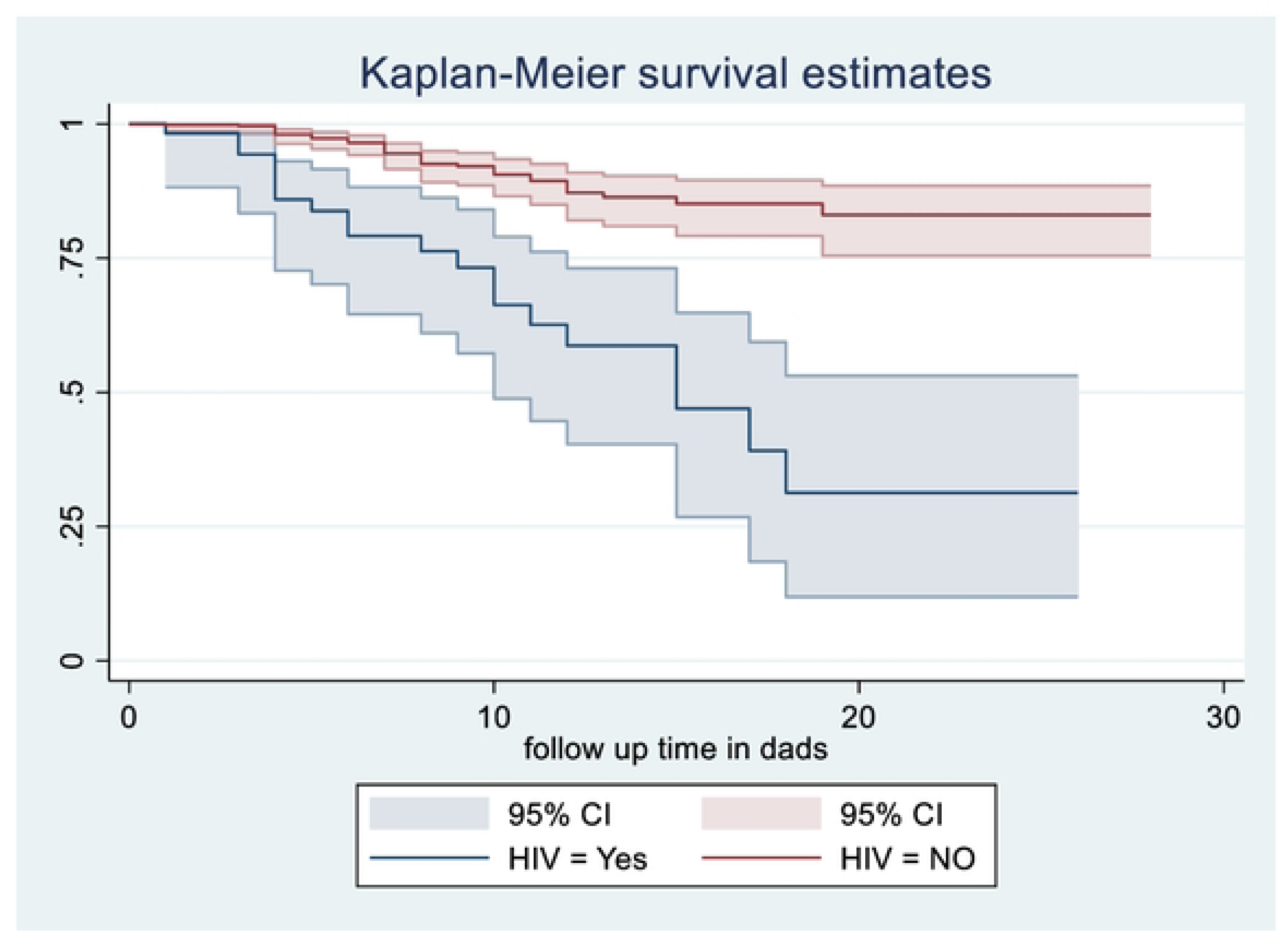
Kaplan-Meir survival estimate among CHD neonates admitted to NICU at public hospitals in Amhara Region with the category of Maternal HIV and without from January 1, 2018-December 30, 2022.

### Factor associated with Incidence of neonatal mortality with Admitted to Congenital heart disease

Cox proportional hazard regression model was used to analyze the relationship between variables and their effect on the recovery time. Bivariate analysis was performed using cox proportional hazard regression to identify which variable had an association with incidence of neonatal mortality to congenital heart disease. Variables like gestational diabetic multiuse (GDM), home delivery, others congenital malformation, sepsis, Airway resuscitation needed during delivery, cyanotic congenital heart disease, were statistically significant with a p-value of 0.25 or less than are predictors of neonatal mortality with congenital heart disease. Therefore, all covariant which had an association with the outcome variable with a p-value of 0.25 or less were entered into the multivariate model.

Multiple cox regressions model was performed for those identified as independent factors in bivariate analysis. Therefore, variables like gestational diabetic multiuse (GDM), home delivery, others congenital malformation, sepsis, Air way resuscitation needed during delivery, and cyanotic congenital heart disease were found to be statistically significant predictors with a p-value of 0.05.

In multivariate analysis, the result shows that the neonates with congenital heart disease who also had neonatal sepsis died 2.24 (AHR: 2.24 (95% CI;-1.18-4.23) times more frequently than those with congenital heart disease who did not have neonatal sepsis (AHR: 2.24 (95% CI;-1.18-4.23), Neonates with cyanotic congenital heart disease had a(AHR 3.49 (95% CI: 1.93-6.28) times higher mortality rate than those with a cyanotic congenital heart disease. Neonates with congenital heart disease were 1.9 times more likely to die at home delivery than those who were delivered in health institution (AHR: 1.9 (95% CI: 1.06-3.6))

The death rate for CHD neonates with a history of maternal GDM had (AHR 1.94(95%CI 1.04-3.61) time’s higher mortality rate than those with congenital heart disease who did not have maternal History of GDM (AHR 1.94 (95% CI 1.04-3.61).Neonates having another congenital malformations. (AHR 2.49(1.33-4.67) times increased the risk of neonatal death compared to not having another one. (AHR 2.49(1.33-4.67). The risk of death among CHD neonates who required airway resuscitation during delivery increased by 57% compared to those who did have (AHR: 0.43 (95%;-0.18-0.98)) (show table 7).

**Table 7:**
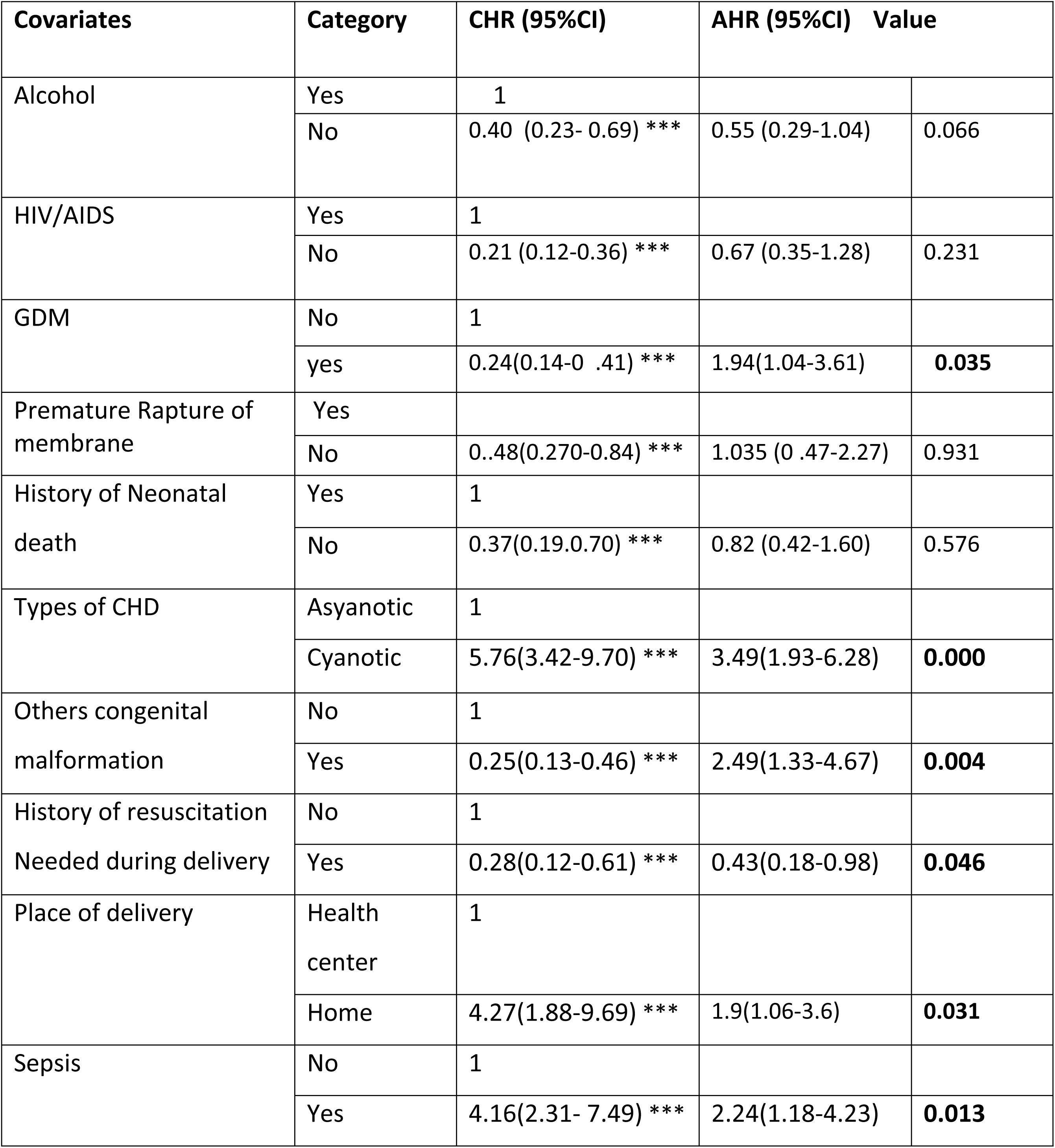
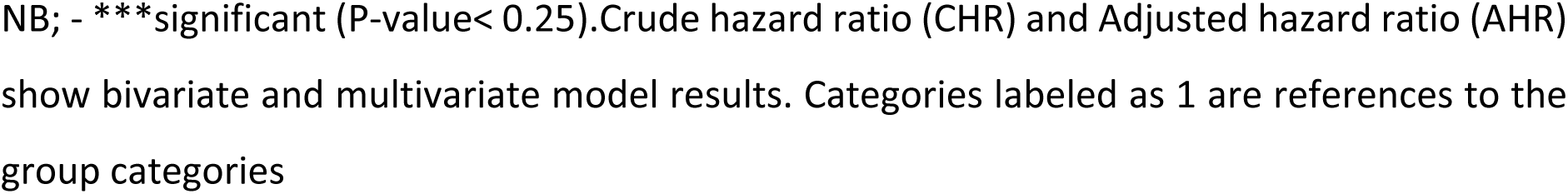
Bivariate & multivariate Cox proportional hazard regression outputs of Congenital Heart disease neonates admitted to NICU at Amhara Region public comprehensives specialized hospitals from Jannuary1, 2018-December 30/2022 in Ethiopia.

### Test of proportional hazard assumption by Schoenfeld’s residuals

Testing the proportional hazard assumption is vital for the interpretation and use of fitted proportional hazard models. Therefore, in this study goodness-of-fit (GOF) particularly the Schoenfeld residuals proportional hazard assumption test for the individual covariates and the global test was used. If P-Value < 0.05, then the proportional hazard assumption is violated. Each covariate (P-Value > 0.05) and all of the covariates simultaneously (Global test for Cox proportional hazard P-Value=0.6337> 0.05) met the proportional hazard assumption (Show table 8).

**Table 8:**
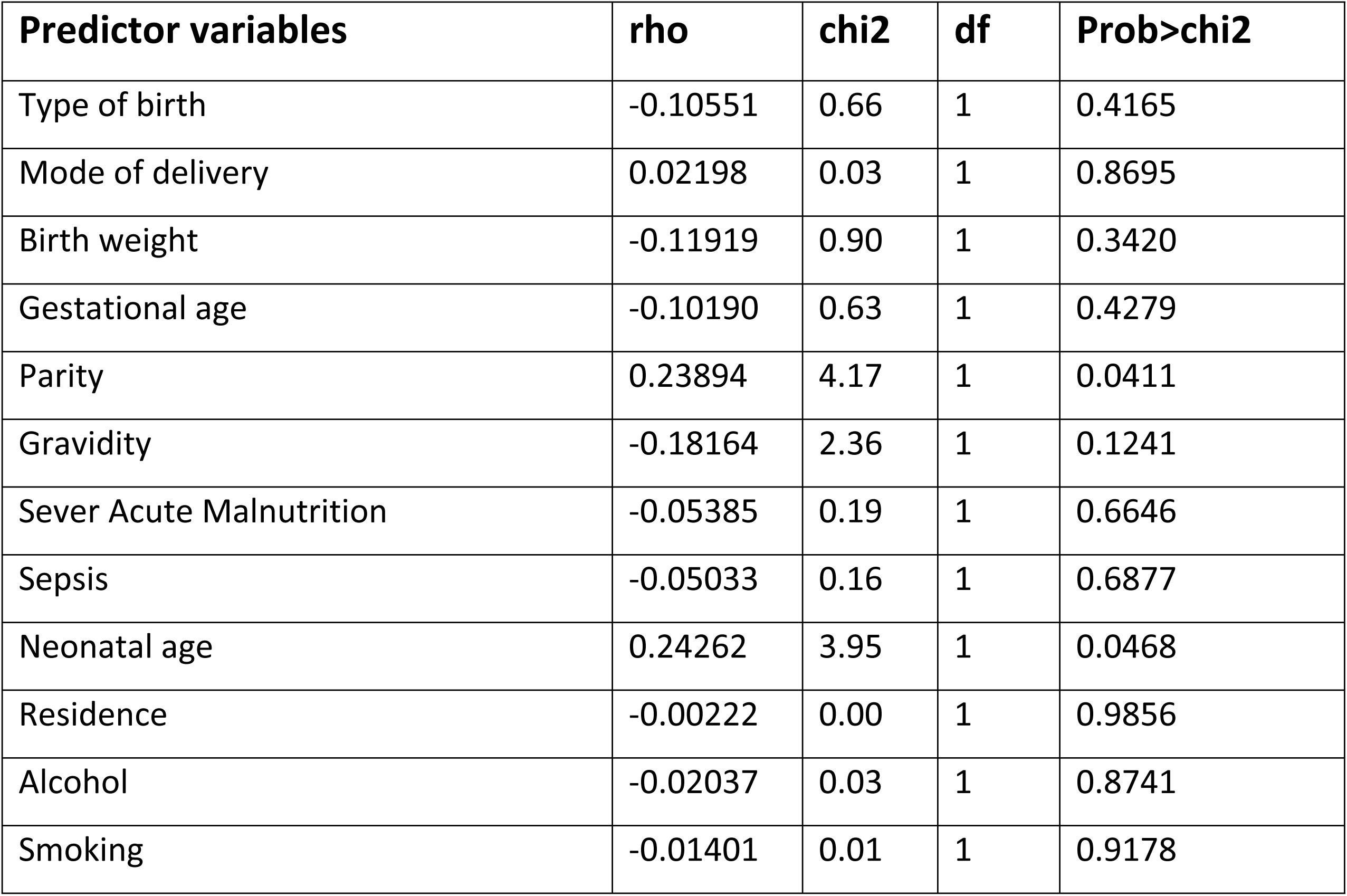
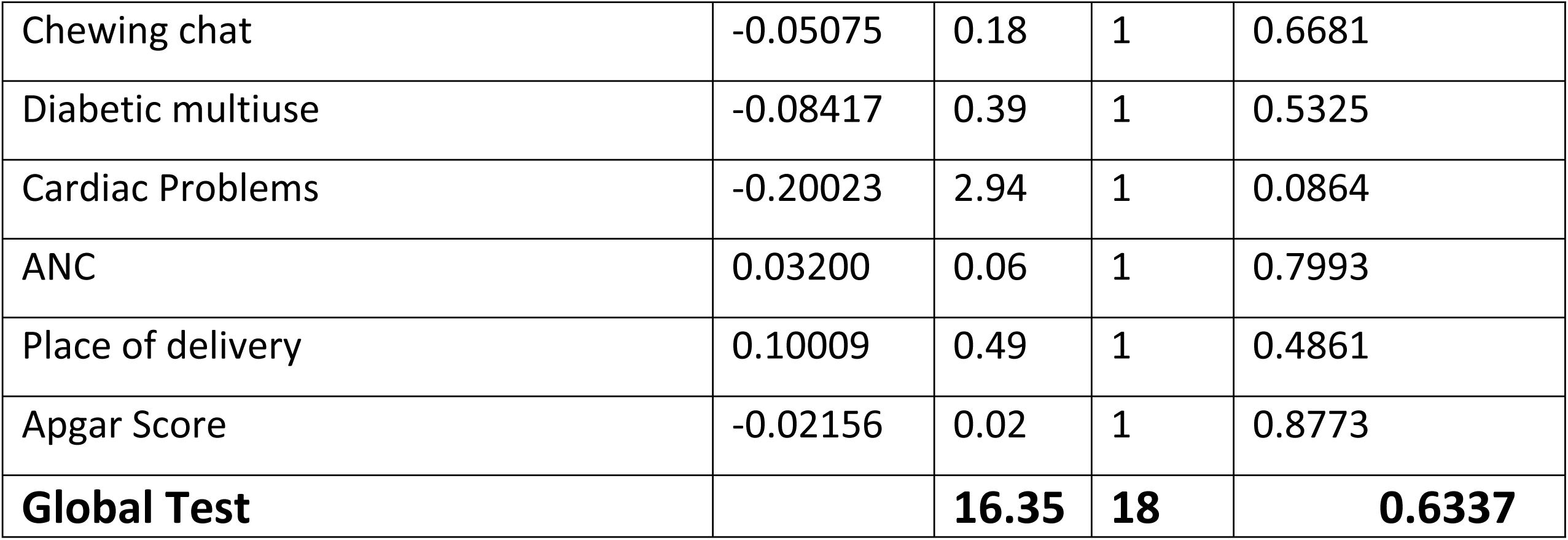
Schoenfeld Residuals test for Cox-Proportional Hazard Regression model assumption (estat, phtest) of each variable for neonates admitted in Amhara Region comprehensives specialized Hospitals in Ethiopia, January 1st, 2018 – December 31th, 2022.

| Predictor variables | rho | chi2 | df | Prob>chi2 |
| --- | --- | --- | --- | --- |
| Type of birth | -0.10551 | 0.66 | 1 | 0.4165 |
| Mode of delivery | 0.02198 | 0.03 | 1 | 0.8695 |
| Birth weight | -0.11919 | 0.90 | 1 | 0.3420 |
| Gestational age | -0.10190 | 0.63 | 1 | 0.4279 |
| Parity | 0.23894 | 4.17 | 1 | 0.0411 |
| Gravidity | -0.18164 | 2.36 | 1 | 0.1241 |
| Sever Acute Malnutrition | -0.05385 | 0.19 | 1 | 0.6646 |
| Sepsis | -0.05033 | 0.16 | 1 | 0.6877 |
| Neonatal age | 0.24262 | 3.95 | 1 | 0.0468 |
| Residence | -0.00222 | 0.00 | 1 | 0.9856 |
| Alcohol | -0.02037 | 0.03 | 1 | 0.8741 |
| Smoking | -0.01401 | 0.01 | 1 | 0.9178 |
| Chewing chat | -0.05075 | 0.18 | 1 | 0.6681 |
| Diabetic multiuse | -0.08417 | 0.39 | 1 | 0.5325 |
| Cardiac Problems | -0.20023 | 2.94 | 1 | 0.0864 |
| ANC | 0.03200 | 0.06 | 1 | 0.7993 |
| Place of delivery | 0.10009 | 0.49 | 1 | 0.4861 |
| Apgar Score | -0.02156 | 0.02 | 1 | 0.8773 |
| **Global Test** |  | **16.35** | **18** | **0.6337** |

### Cox-Snell residuals Test

Cox-Snell residuals test is used to assess the overall goodness of fit in survival models. It is done graphically with the usual graphs of Cox-Snell, and it is observed that residuals from a correctly fitted model follow exponential unite distribution along the 45 degree of slope (baseline). It is used to identify extreme observations that need additional investigation. In this study, the hazard function follows the 45-degrees line closed to the baseline. It showed that the multivariate cox regression model is fitted for the analysis and interpretation of the results of this study (fig.8)

**Figure 8:**
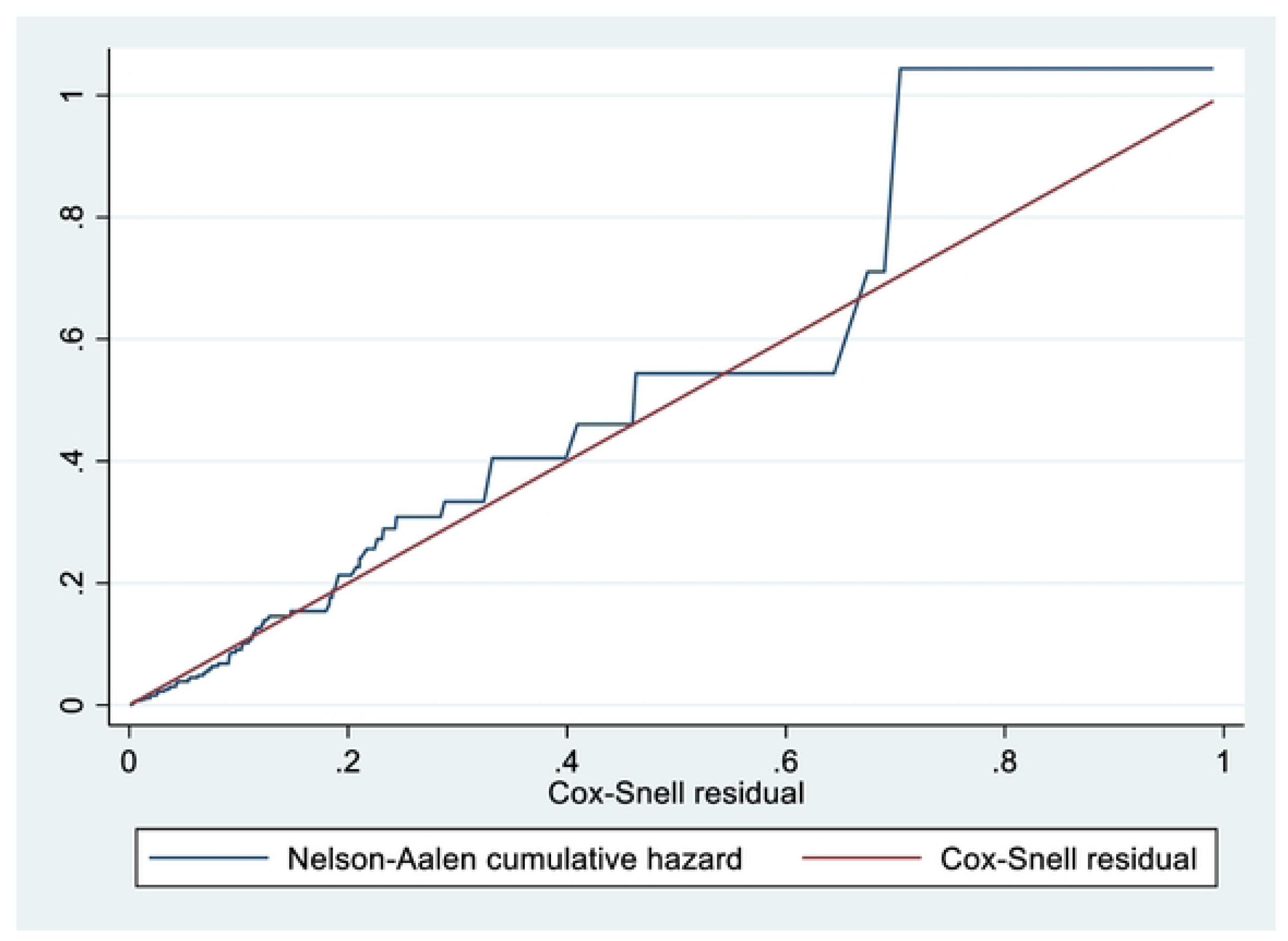
Cox-Snell residual cumulative hazard graph on neonates admitted in NICU with congenital heart disease Amhara Region at Comprehensives Specialized Hospitals in Ethiopia, January 1st, 2018 December 31th, 2022 in Ethiopia.

## Discussion

Incidence of mortality among neonates with congenital heart disease was 11.9 (95% CI; - 0.009-0.015) per 1000 persons of observation which was in line with a study conducted, USA neonatal mortality (0.37/1000%)[48], in gingdao china (9.38/1000%),[49],south china was approximately 11.1 per 1,000 live births [50] and Sri lanka 1.56/1000 live birth[51], The possible reason could be due to the study were conducted at the hospital level, in which different complicated neonatal cases can be referred from different corners of the region, so due to a high number of complicated cases the mortality could be increased.

However, it was higher than the study conducted in Europe (0.7%/1000) [52] & Spain (4.58/1000). [53], This variation might be due to the NICU physical environment difference. For example, in developed countries, there is a dedicated ward space and enough neonatal beds even though the distance between NICU and the labor ward is closely constructed. The other possible reason might be the ward of those developed nations might be staffed with health workers with specialty training and skills like neonatologists, pediatric surgeon & nursing specialties than sub saran Africa including Ethiopia. Another possible explanation might be also a difference in the health care utilization of mothers due to discrepancies in sociocultural factors.

On the contrary finding lower than the study conduct in Sweden northern European country 17/1000(95% CI, 16.8–18.6)[11] and brazil/ (85%/1, 0000), births[54]this might be due to differences in length of follow-up time, study period, study area, and study design

A multivariate analysis found that neonates with congenital heart disease who also had neonatal sepsis died 2.24 (AHR: 2.24 (95% CI;-1.18-4.23) times more frequently than those with congenital heart disease who did not have neonatal sepsis (AHR: 2.24 (95% CI;-1.18-4.23), This was in line with a study conducted in, TASH, Addis Ababa, Ethiopia, were 4.4 times more likely to dies [AHR=4.418 (95%CI: 1.617-12.072)].[55] Northern and south western Ethiopia [35, 55–57]and united states of America.[58].this discrepancy may be study area, Congenital heart disease neonates have decreased immune function because of the low level of some immune cells such as immunoglobulin G resulting from the low transfer of maternal immunoglobulin G and predispose to varies health problems like malnutrition, developmental delays and infection and neonates with CHD with increased pulmonary blood flow thus presents with pneumonia and congestive cardiac failure (CCF) amongst other features, neonates with congenital heart diseases are more vulnerable to infections following the fact that they had an anatomical defect that causes hemodynamic disturbance of lung circulation and mucosal edema finally leads to decreased lung compliance and develop infections. So preventing neonatal sepsis for neonate with congenital heart disease is important to decrease neonatal mortality and helps to implement SDG

Neonates with cyanotic congenital heart disease had a 3.49 (95% CI: 1.93-6.28) times higher mortality rate than those with asyanotic congenital heart disease. This result was consistent with a study done in Atlanta, Georgia (69) Spain (59), this difference may be due to differences in model analysis, neonates are at risk of having serious complications within the first few days or weeks of life, often require emergency care and is silent in fetal life B/C right-to-left shunting across the foramen ovals and ductus arteriosus. in the presence of pulmonary outflow tract obstruction or persistent pulmonary hypertension of the newborn. Which could increase the risk of dying than Acyanotic congenital heart disease

Neonates with congenital heart disease were 1.9 times more likely to die at home delivery than those who were delivered in health institution (AHR: 1.9 (95% CI: 1.06-3.6)).This finding was supported by studies in Iran, Sub-suharan Africa(56),Ethiopia[30], in Ethiopia due to financial problems, lack of knowledge, preference of traditional medicines for a sick neonates, poor roads. Poor health facility readiness, the poor economy of families, lack of ambulance, cultural and traditional beliefs, and unprofessional behaviors which were the major barriers hindering the uptake of delivery of health care facility. This finding indicates that programs and policies should focus on the strengthening of the quality of care in maternal and child health.so improving the quality of care could be important to avoid the negative attitude of mothers towards the services provided by health institutions,

The death rate for CHD neonates with a history of maternal GDM had (AHR 1.94(95%CI 1.04-3.61) time’s higher mortality rate than those with congenital heart disease who did not have maternal History of GDM (AHR 1.94 (95% CI 1.04-3.61). This study finding is consistent with the study conducted at in Ethiopia Addis Ababa hospitals (47). This might be GDM related to the complications Protected due to diabetes mellitus during pregnancy because diabetes mellitus can result in hypoglycemia, hypocalcaemia, respiratory distress, growth restriction, polycythemia, increased magnesium amount, respiratory distress, congenital anomalies, and excessively increased amount of bilirubin

According to this study. Neonates having another congenital malformations. (AHR 2.49(1.33-4.67) times increased the risk of neonatal death compared to not having another one.(AHR 2.49(1.33-4.67).This finding is supported by studies done by TASH Addis Ababa Ethiopia ([AHR=2.416 (95%CI: 1.367-4.264)] (76) and higher the study conduct in Finland (7 to 50% of the neonates with congenital heart disease with other congenital malformation a greater risk of comorbidity and mortality [59] This discrepancy might be due to difference in methodology, lifestyle, economic status, access to health care facilities, and level of education of the general public. may have contributed to increased incidence of risk of mortality to CHD in Ethiopia. Furthermore, poor access to health care facilities in our country, accompanies many patients to seek alternative treatments such as, using herbal remedies, prayers and rituals that encountered a delay in care which further complicating the disease process,

The risk of death among CHD neonates who required airway resuscitation during delivery increased by 57% compared to those who did have (AHR: 0.43 (95%;-0.18-0.98)),this finding is also higher the study conducted in Belgium is 36%[60] The possible reason for the observed discrepancy might be due to the difference in sample size, study setting, the severity of the disease, and the methodology of studies.

Transition from fetal to postnatal life is a complex process. Even in the absence of congenital heart disease, about 4–10% of newborns require some form of assistance in the delivery room. Neonates with complex congenital heart disease should be expected to require significant intervention and thus the resuscitation team must be well prepared for such a delivery.

## Conclusion

The rate of mortality among hospitalized congenital heart disease neonates in this study was high as compared to findings of other studies, in this study the overall incidence of congenital heart disease neonatal mortality was 11.9/ per 1000 day observation, congenital heart disease neonates from sepsis, Home delivery, maternal history of GDM, other congenital malformation, Asyanotic CHD and neonatal resuscitation need careful observation and early intervention of cases is crucial to prevent the potential CHD related mortality, Furthermore, strengthening the health care delivery system to satisfy the need of the patients should get due attention to reduce the incidence of mortality from CHD cases

### Strength and Limitations

#### Strength

The study area covers Amara region Referral Hospital. As a result, its generalizability to all hospitals of Amhara region in may be possible. This study was conducted by using five consecutive years of admission and Study participants were included from different healthcare institutions, which result in to increase in its representatives.

#### Limitations

Some critical covariates obtain from the mother were missing. These might be significant predictors of neonatal mortality such educational status, occupational status, nutritional status, economic status and did not include biomedical profiles and medication-related variables which could be potential predictors of congenital heart disease neonatal mortality.in addition to the study was retrospective cohort study design

#### Recommendations

Even though quality of health care service expanded, especially for maternal and newborn care, neonatal mortality is still a concerning issue in all Ethiopia’s study areas. Based on the findings of this study, I forwarded the following recommendations.

#### Policy Makers and Local leaders

As already known, currently, there is only one surgical treatment center for neonates with congenital heart disease at TASH. FOMH should expand the surgical treatment centers other than TASH because almost all cases of congenital heart disease were referral cases and the majority were died due to delay in surgical intervention and delaying reaching care. FMOH should also incorporate the standard of care for neonates diagnosed with congenital heart disease in the national guidelines. The accessibility advanced care center and quality of surgical interventions also need emphasis to reduce incidence of mortality neonates with heart disease. Collaborative and advanced care should be considered for neonates with congenital heart disease.

#### Amhara Regional Health Bureau

Amhara regional health bureau had better strengthen the treatment strategies of neonates diagnosed with congenital heart disease by supplying the equipment needed for intervention like mechanical ventilation, Suction machine Echocardiography and isolated Operation room for timely surgical intervention and should make availability of total parenteral nutrition in the hospitals

#### Health Professionals and Health Institutions

To increase health facility delivery, health care providers and health care facilities strengthening raising women’s awareness on the benefits of delivering in health facility, male involvement in the use of maternal health services, increasing women decision-making power, addressing common barriers of lack of transport, and compensations for transport expenses to alleviate the cost of transport and health education focusing on the potential threats of delivering at home at recommended in the health care providers

Health care provider had screened and tasted for GDM typically between 24-28 weeks of gestation and increase awareness of balanced diet

All health care providers had insures that all babies receive essential newborn care (ENC), including appropriate cord care, early initiation and exclusive breastfeeding, maintenance of warmth, and prompt treatment with antibiotics and who are working in labor and delivery ward should be give attention for mothers those have antenatal, intrapartum fever and refer the neonates to NICU for early initiation of management

Pregnant mothers should attend regular ANC follow-up and early diagnosis of congenital heart disease is vital to improve the quality of care provided for newborns

#### To future researchers

Upcoming researchers shall conduct a longitudinal prospective cohort study to incorporate impossible variables in retrospective study like maternal socio-economic, genetic, biomedical profiles and medication-related variables and environmental factors and a qualitative study should be conducted to explore the possible psychosocial support for mothers who lost their babies due to congenital heart disease.

## Data Availability

All data were included in the manuscript and we will submitting as soon as you request

## Acknowledgments

First, we want to express our heartfelt gratitude to Debre Berhan University Asrat Woldeyes Health Science Campus School of Nursing & Midwifery for giving valuable support. Secondly, the deepest gratitude to all Pre-Publication Support Service (PRESS) of PLOS ONE, the staff member of amhara region comprehensives specialized hospitals, study participants and the data collectors their cooperation in this study.

## Supporting information

<u>S1 IRB letter</u>

<u>S2 Plos one studies clinical checklist</u>

<u>S3 STROBE-statement checklist</u>

## Authors’ contributions

**Conceptualization:** Abatwoy Ayfokru.

**Data curation:** Abatwoy Ayfokru.

**Formal analysis:** Abatwoy Ayfokru.

**Investigation:** Abatwoy Ayfokru, Sisay Shewasinad, Fuad Ahmed, Mitiku Tefera,Emawaysh Getaneh and Genet Nigusie

**Methodology:** Abatwoy Ayfokru, Sisay Shewasinad, Fuad Ahmed, Mitiku Tefera,Emawaysh Getaneh and Genet Nigusie.

**Project administration:** Abatwoy Ayfokru, Sisay Shewasinad, Fuad Ahmed, Mitiku Tefera,Emawaysh Getaneh and Genet Nigusie.

**Resources:** Abatwoy Ayfokru, Sisay Shewasinad, Fuad Ahmed, Mitiku Tefera,Emawaysh Getaneh and Genet Nigusie.

**Software:** Abatwoy Ayfokru, Sisay Shewasinad, Fuad Ahmed, Mitiku Tefera,Emawaysh Getaneh and Genet Nigusie.

**Supervision:** Abatwoy Ayfokru, Sisay Shewasinad, Fuad Ahmed, Mitiku Tefera,Emawaysh Getaneh and Genet Nigusie.

**Validation:** Abatwoy Ayfokru, Sisay Shewasinad, Fuad Ahmed, Mitiku Tefera,Emawaysh Getaneh and Genet Nigusie.

**Visualization:** Abatwoy Ayfokru, Sisay Shewasinad, Fuad Ahmed, Mitiku Tefera,Emawaysh Getaneh and Genet Nigusie.

**Writing – original draft:** Abatwoy Ayfokru, Sisay Shewasinad, Fuad Ahmed, Mitiku Tefera,Emawaysh Getaneh and Genet Nigusie.

**Writing – review & editing:** Abatwoy Ayfokru, Sisay Shewasinad, Fuad Ahmed, Mitiku Tefera,Emawaysh Getaneh and Genet Nigusie.

### Acronyms and abbreviation

ANC: Antenatal Care
AHR: Adjusted Hazard Ratios
ARHB: Amhara Regional Health Bureau
CHD: Congenital Heart Disease
CHR: Crude hazard ratio
CI: Confidence interval
CPH: Cox proportional Hazard Regression
GDM: Gestational Diabetes Mellitus
LIMIC: Low Income and Middle Income Countries
MOH: Ministry of Health
NICU: Neonatal intensive care units
SD: Standard deviation
TASH: Tikur Anbesa specialized Hospital
TOF: Tetralogy of Fallot
USA: United States of America

